# Exploring the experiences and support recommendations of autistic adults drinking alcohol

**DOI:** 10.64898/2026.08.12.26360339

**Authors:** Stephanie Page, Kayleigh E Easey, Felicity Sedgewick, Dheeraj Rai, Evie Stergiakouli

## Abstract

Autistic individuals may be at an increased risk of hazardous drinking compared to non-autistic counterparts: potential motivations include facilitated social interactions and self-medication of co-occurring difficulties, and possible risk factors include being older and female. However, research remains limited and centred around clinical samples. Given the diversity of the autistic community, it is important to understand the intricacies of alcohol use to inform appropriate support. Eighteen autistic adults took part in semi-structured interviews about their drinking experiences. Data were analysed using reflexive thematic analysis. Three main themes were created (“Autistic experiences”, “Managing expectations and coping by drinking” and “Recommendations for support”). “Autistic experiences” was used to denote the ways in which participants described their own autistic features influenced their relationship with alcohol. “Managing expectations and coping by drinking” was chosen to reflect the pressures felt by participants to show up in social relationships and the co-occurring difficulties many of them managed using alcohol. Therapeutic preferences for alcohol services were captured under “Recommendations for support”. As expected, participants used alcohol to facilitate social interactions and self-medicate. However, additional nuances uncovered may provide clinical utility and highlight the need for further research in other demographics within the community.

## Introduction

Results from quantitative studies suggest a complicated relationship between autism and alcohol use. Previous findings suggest that autistic people are less likely to consume alcohol hazardously and become dependent than their non-autistic counterparts.^1–9^ In a large Swedish cohort study (N=20,863), individuals with a proxy diagnosis for autism were less likely to develop alcohol use disorders than twins who either did not have the diagnosis or had the diagnosis with co-occurring attention deficit hyperactivity disorder (ADHD) or learning difficulties.^3^ Conversely, emerging studies suggest that autistic people might actually be more likely to drink hazardously than non-autistic people.^10,11^ Another Swedish cohort study (N=123,453) found that compared to their non-autistic relatives, N=26,986 autistic individuals without learning difficulties were twice as likely to report alcohol related problems.^10^ Further, regression analyses involving a community sample of 237 autistic adults revealed a U-shaped relationship between increased autistic traits and hazardous drinking and teetotalism.^12^ These findings suggest that certain members of the community may struggle with their drinking, yet it is unclear what drives this relationship in some autistic people but not others.

Preliminary studies have unearthed a few motivations for alcohol use in autistic people. In their online survey involving 507 autistic adults, Brosnan and Adams found that heavy episodic drinkers were more likely than lighter drinkers to endorse the notion that drinking alcohol when autistic makes communication easier.^13^ This was reflected in a narrative systematic review of 22 studies, in which the authors theorised that the positive reinforcement of facilitated social interactions was a driving factor for alcohol use in autistic people.^14^ Other potential motivations suggested in this review included self-medication of overwhelm and overstimulation, in addition to increased confidence, comfort and inclusion.^14^ However, the studies in which these motivations were identified included clinical samples only (i.e. autistic outpatients who were accessing psychiatric services). Given that research remains limited and the community is diverse generally, further investigation is warranted to understand potential nuances of the issue in non-clinical samples and inform appropriate support.

Qualitative pattern-based approaches can complement quantitative methods by illuminating details from lived experiences that would otherwise go undetected.^15^ Some qualitative work on alcohol use in the autistic community exists, but studies remain limited. Rothman et al. conducted 40 semi-structured interviews with formally diagnosed autistic individuals between the ages of 16-20 about their motivations for drinking or abstaining from alcohol^16^ Using content-based analysis, the authors identified three themes. The first theme, “Deliberate about alcohol use decisions”, highlighted the deliberative decision-making participants reported around drinking, which included doing their own research about the potential effects of alcohol and fear of becoming inebriated. The second theme, “Reasons for not drinking”, encompassed the reasons that participants avoided drinking, including fear of addiction, unpleasant taste, potential medication interactions, potentially exacerbated anxiety and fearing the inability to mask due to disinhibition. The final theme, “Reasons for alcohol use”, denoted reasons that participants chose to drink. These included having to mask less in social settings, feeling less irritable or bored, managing sensory input better, feeling more socially motivated and accepted, and feeling like they coped with their problems better.

Autistic adults over the age of 20 have also reported social facilitation as a motivator for drinking in qualitative studies. Kronenberg et al. interviewed 12 adult outpatients receiving substance treatment who were formally diagnosed with autism and substance use disorder (SUD) about the consequences of their substance use.^17^ One participant found it easier to explain themselves when they had consumed alcohol, while another said they would not know how to socialise without drinking and was therefore reluctant to give up completely. This is reflected in qualitative findings from Clarke et al., in which eight participants were recruited from autism or drug and alcohol services and interviewed about the initiation and maintenance of their substance use (including alcohol).^18^ Further, both Kronenberg et al. and Clarke et al. also found that participants used alcohol to self-medicate for various reasons, including the stress of parenthood, boredom, co-occurring depression, anxiety and/or sleep issues.^18,19^

Evidently, autistic people drink for a variety of reasons, which is unsurprising given the diversity of the community generally. However, qualitative research in this area remains limited, both in terms of its existence and by the use of predominantly clinical samples. Alcohol use is a prerequisite for hazardous drinking and alcohol dependence. It is therefore vital that the nuances of this are better understood in the autistic community to help our understanding to how autistic people may transition from alcohol use into hazardous drinking and/or dependence. Furthermore, no qualitative studies appear to ask autistic people directly about their preferences for clinical support regarding alcohol use. Autistic people have expressed a desire to be directly involved in research and asking about this provides a platform from which the community could voice their needs.^20^

This study aims address the lack of general population samples in previous studies and explore clinical support recommendations made by autistic people in relation to alcohol use. In doing so, the paper aims to answer the following research questions:

*RQ1: What are the experiences of autistic people around drinking alcohol from adolescence to adulthood?*

*RQ2: What recommendations do autistic people have around support for struggling with their drinking?*

## 6.2 Methods

### Participants

Eighteen participants took part in semi-structured interviews. Participants were eligible if they were over the age of 18, fluent English speakers, and had a formal diagnosis of autism from a clinician or would self-describe as being autistic. Participant demographics are given in Table 1.

**Table 1.**
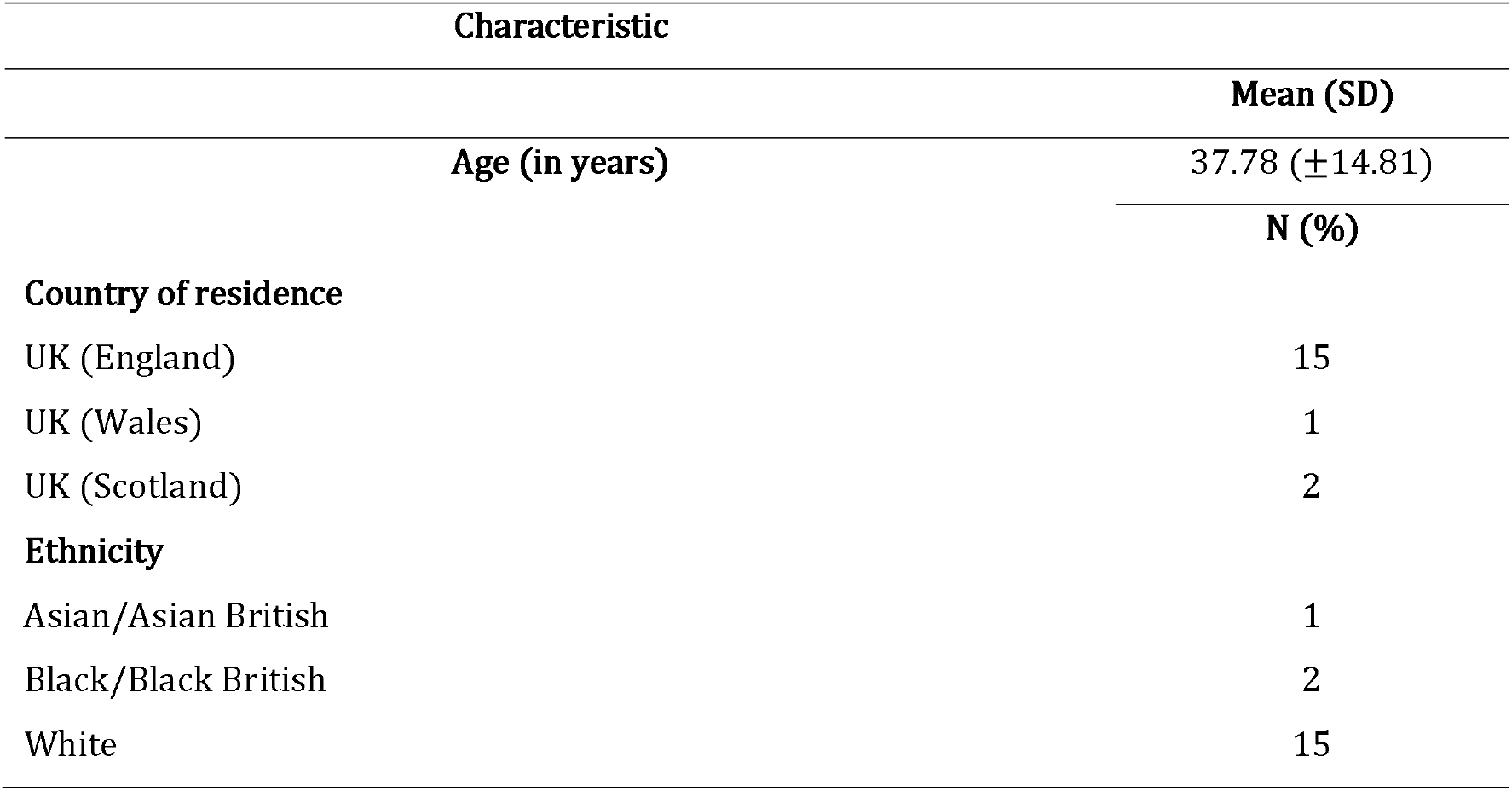
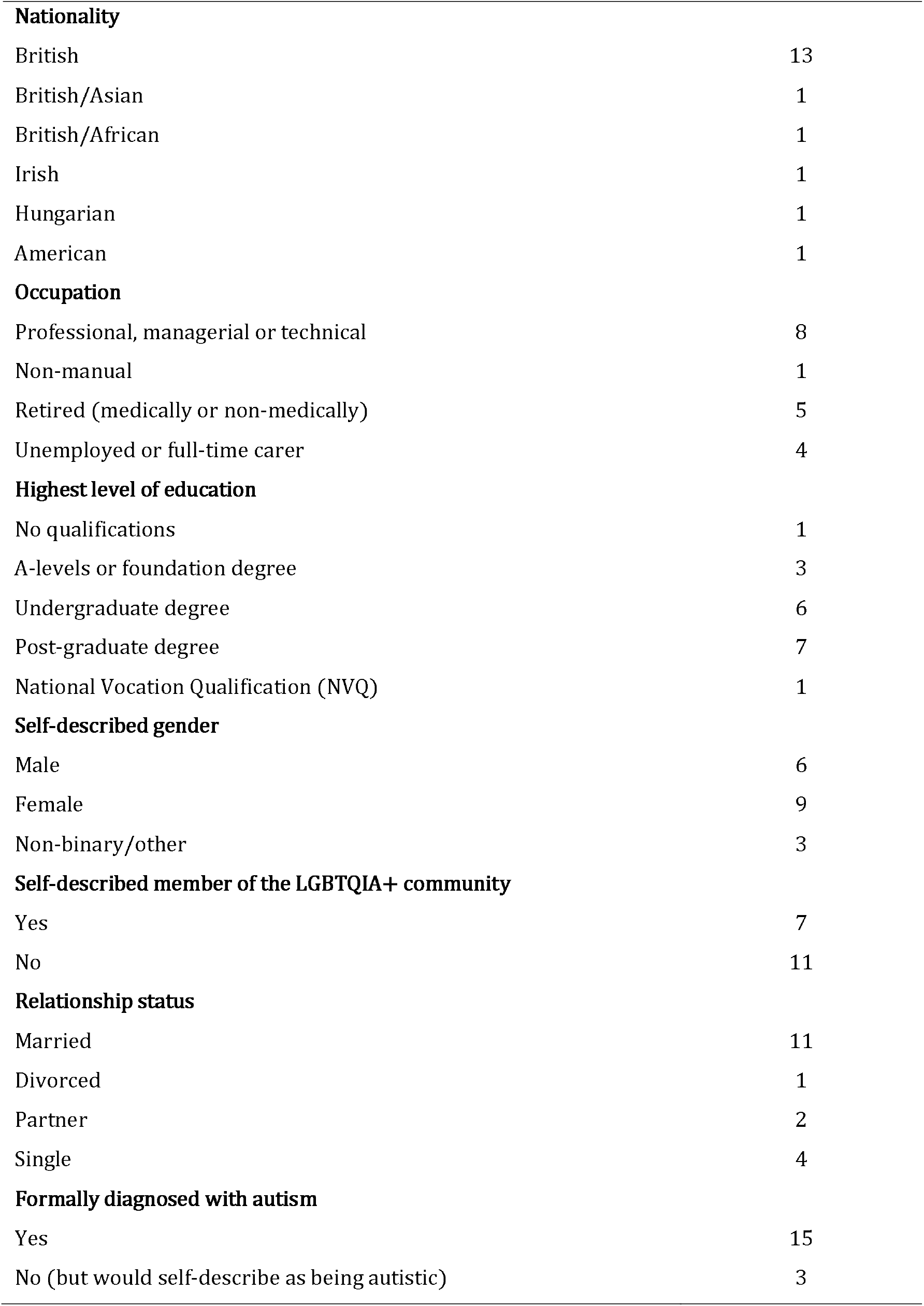
Participant demographics (data collected during screening calls).

| Characteristic |  |
| --- | --- |
|  | Mean (SD) |
| Age (in years) | 37.78 (±14.81) |
|  | N (%) |
| **Country of residence** |  |
| UK (England) | 15 |
| UK (Wales) | 1 |
| UK (Scotland) | 2 |
| **Ethnicity** |  |
| Asian/Asian British | 1 |
| Black/Black British | 2 |
| White | 15 |
| **Nationality** |  |
| British | 13 |
| British/Asian | 1 |
| British/African | 1 |
| Irish | 1 |
| Hungarian | 1 |
| American | 1 |
| **Occupation** |  |
| Professional, managerial or technical | 8 |
| Non-manual | 1 |
| Retired (medically or non-medically) | 5 |
| Unemployed or full-time carer | 4 |
| **Highest level of education** |  |
| No qualifications | 1 |
| A-levels or foundation degree | 3 |
| Undergraduate degree | 6 |
| Post-graduate degree | 7 |
| National Vocation Qualification (NVQ) | 1 |
| **Self-described gender** |  |
| Male | 6 |
| Female | 9 |
| Non-binary/other | 3 |
| **Self-described member of the LGBTQIA+ community** |  |
| Yes | 7 |
| No | 11 |
| **Relationship status** |  |
| Married | 11 |
| Divorced | 1 |
| Partner | 2 |
| Single | 4 |
| **Formally diagnosed with autism** |  |
| Yes | 15 |
| No (but would self-describe as being autistic) | 3 |

**Table 1.** Quotes illustrating subthemes of “Autistic experiences”.

| Subtheme | Quote | Participant # |
| --- | --- | --- |
| **Diagnostic criteria and alcohol** |  |  |
| **Social communication differences** | “It helped me feel more comfortable because when people get drunk, they often become more honest.” | 9 |
|  | “Later, it became a strategy to help me fit in. I felt my communication was better - I felt I could understand people more, or rather, that they could understand me better when I had been drinking.” | 8 |
| **Restrictive and repetitive behaviours and interests** | “...once it became part of my routine, it felt like something was missing if I didn’t do it | 1 |
| **Sensory differences** | “I think it helps to dull things. If I'm in an environment like a pub, a party, or a social event, having a drink helps to ease the anxiety caused by the noise, the smells, and everything else. It kind of numbs everything on all levels. So, yes, it definitely helps.” | 14 |
| **Masking** | “I couldn’t disassociate anymore. Alcohol was my quickest and easiest way of going into stasis, so I could wake up the next day, put my mask on, and go to work.” | 11 |
| **Risk aversion** | “I’ve always stayed away from alcohol. I’ve never done drugs in my entire life—not even a cigarette. That’s more in line with my tendency to opt out of things—maybe over-worrying, maybe just avoiding risks.” | 4 |

### Ethical approval

Ethical approval was granted by the Faculty of Health Sciences Research Ethics Committee (FREC) at the University of Bristol (Ref: 18949).

### Procedure

The study was advertised virtually via the Autistica research network. Participants were invited to attend a short screening call via Microsoft Teams in which they would be asked demographic questions. These questions enquired about participant age, country of residence, ethnicity, nationality, occupation, education, self-described gender, self-described member of the LGBTQIA+ community, relationship status and formal diagnosis of autism or self-described autistic person. They were also asked about their motivations for applying. Participants were made aware that they would need to ideally have their camera on for this, as a form of verification of the authenticity of their participation.^21^ For the main interview, participants had a choice of attending via Microsoft Teams with their camera on or off or could respond to questions via email if preferred to improve study accessibility (although none of the participants chose to have their camera off).^22^ One participant responded to questions via email, but this was due to technical difficulties.

A total of 201 individuals expressed an interest in taking part in the study. Of this group, the first 93 available individuals were invited to attend short screening calls on three separate days when S.P. was available. After finishing the screening calls, 20 participants were invited to interview with S.P. Participants were selected based on their motivations for applying to the study and differing demographics to help reflect the diversity of the autistic community. Two participants dropped out of the study, which left eighteen participants to complete the semi-structured interviews. Participants were provided with an information sheet prior to booking an interview to help them decide whether they would like to take part. This is so that they understood the purpose of the research and were made aware they could withdraw at any time so they could provide their informed consent.

Interviews ranged from 20 minutes to 65 minutes long (mean=41.56 minutes, SD=15.34 minutes). For accessibility, participants were given the option of seeing the participant version of the topic guide prior to the interview.^23^ The interview topic guide was developed through consultations with an advisory group of five autistic adults (also recruited via the Autistica network). Group members gave feedback on language used and which topics relating to alcohol use that they felt were important to cover. Following approval from the advisory group, the topic guide was then used in the interviews. Advisory group members were reimbursed at £25 per hour for their time. Participants received a £25 Amazon voucher once they had completed the interview. Interviews were recorded and automatically transcribed using the transcription feature of Microsoft Teams. Transcripts downloaded from Microsoft Teams were then checked by S.P. against the recording. Participants were given two weeks to request that their interview responses be deleted before transcripts were anonymised.

### Data analysis

Interview data were analysed using RTA. This is because research in this area remains very limited, and the authors wanted to encourage the appearance of unknown nuances rather than categorising data according to pre-existing theoretical constructs (as with codebook or coding reliability TA).^24^ The epistemological approach drawn upon in this paper was constructivist, as the authors generated meaningful overarching patterns that they had interpreted subjectively from participant experiences.^25^ In line with this, data were coded inductively; no pre-defined codes were used, but rather new codes generated by authors throughout the analytic process. These were based on what felt relevant to the research questions, but also what appeared important to each participant individually. Deductive coding was not used in this study, as the authors felt it this would not align the chosen analytical method (given that RTA is designed around the generation of novel codes).^26^ Coding was also semantic in that the codes generated were intended to reflect what participants had said explicitly. This was for two reasons; the first was that the authors wanted to stay as true to participants experiences as possible. This is because part of the study rationale was to provide a platform for autistic people to express their needs in relation to alcohol use. The second reason for semantic coding was that the interview schedule was designed to gather information directly relevant to the RQs 1 and 2, so deeper interpretation felt unnecessary. The interpretations in the analysis were predominantly from author one (S.P.). However, author two, F.S. also contributed by acting as a sounding board for S.P. to discuss interpretations and offer alternative perspectives on any meaningful patterns generated. This was to honour the non-positivist nature of RTA by enabling S.P. to interpret the data subjectively while helping them to present findings coherently. It was also to ensure that both authors felt participant experiences had been represented as accurately as possible.

In terms of the analytic process, the authors followed phases one to six as outlined by Braun et al. (known as “1) Familiarisation”, “2) Coding”, “3) Initial theme generation”, “4) Reviewing and developing your themes”, “5) Refining, defining and naming themes” and “6) Producing the report”).^24^ Prior to the first phase, interviews were transcribed by S.P. using the Microsoft Teams transcript function (version 25306.805.4102.7211). S.P. then downloaded transcripts onto Microsoft Word documents and checked them manually against interview recordings to ensure accuracy. S.P. then uploaded finalised transcripts to qualitative software package NVivo (version 14).

In the first phase, S.P. and F.S. familiarised themselves with the data by reading and re-reading transcripts (the transcription process also assisted S.P. in this phase). S.P. and F.S. had informal discussions about what was coming up at this stage, commenting on the richness of the data. They then moved into the second phase. Here, S.P. inductively coded all 18 transcripts by using the coding function in NVivo. F.S. inductively coded nine of the 18 transcripts and then the authors met to discuss codes they had generated and identify duplicate codes that could be collapsed to encompass multiple data segments. To clarify, the collapse of duplicate codes was not the point at which themes were generated, but rather to avoid multiples of the same as so many code labels had been created in phase two. For the third phase, S.P. decided to write all finalised code labels onto individual pieces of paper, as they found it more helpful for organising their thoughts than using NVivo for this step. All pieces of paper were laid out on a large table and S.P. was then able to start grouping code labels together under initial themes. An extra initial theme of “Uncategorised” was created so that unassigned code labels were kept for review as part of the iterative process of theme generation. For phase four, S.P. created a digital mind map of initial themes and presented this to F.S. to discuss interpretations and settle on what both authors felt was a sound thematic snapshot of the data. Codes in the “Uncategorised” initial theme were also reviewed to ascertain whether F.S. felt there was a space for them under new or existing themes. In the fifth phase, S.P. produced Figure 1., a polished version of the mind map created in the previous phase and wrote accompanying theme definitions into a drafted results section of the paper to get a sense of how to order themes. The authors then met to discuss the coherence and flow of the themes presented. Lastly in phase six, S.P. finalised theme reporting by ordering themes, linking subthemes across findings and crafting the overall message of the results.

**Figure 1.**
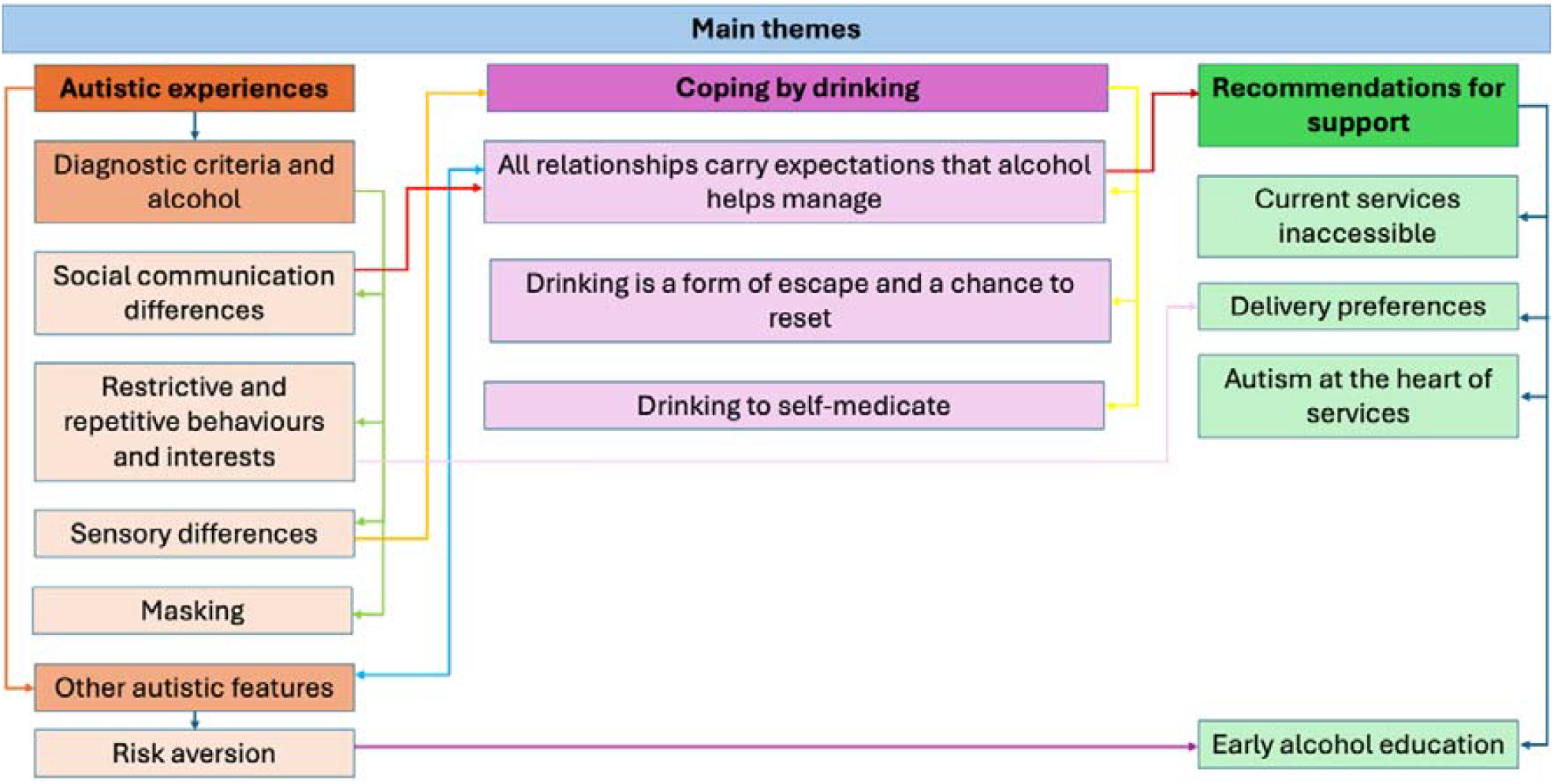
Diagram showing interconnectedness of themes and subthemes

## 6.3 Results

The main themes and subthemes created are given in Figure 1. We generated three main themes: “Autistic experiences”, “Coping by drinking” and “Recommendations for support”, which are discussed in detail below.

### Autistic experiences

Many participants described how their autistic features influenced their relationship with alcohol consumption, resulting in the subthemes of “Diagnostic criteria and alcohol” and “Other autistic features”, reflecting autistic traits specified in the Diagnostic and Statistical Manual of Mental Disorders – Fifth Edition Revision (DSM-5-TR) and additional features known to be prevalent among autistic people (specifically risk aversion).^27^ Supporting quotes are given in Table 1.

### Diagnostic criteria and alcohol

#### Social communication differences

“Persistent deficits in social communications and interaction across multiple contexts” is one of the DSM-5-TR diagnostic criteria for autism.^27^ Hereafter, these will be referred to as social communication differences rather than deficits to avoid insinuating a comparative inferiority of autistic individuals to neurotypical individuals. Social communication differences often make social interactions more challenging for autistic people. Participants described how drinking helped their social interactions by changing their own demeanour (as well as that of others). After drinking, they found it easier to be present, confident, relaxed, understand others, communicate clearly and overthink less. One participant even wished they could experience being the drunk version of themselves all the time because of the social acceptance they felt.

Participants also felt that the people who they were drinking with became more open, interesting to talk to and easier to understand, which made socialising more enjoyable. For many participants, it seemed like drinking alcohol could help them feel more connected to those around them compared to socialising when sober. It is not just autistic people who find socialising easier when drinking alcohol. However, these findings suggest drinking may be particularly helpful for autistic people when socialising and exemplifies how autistic traits can interact with drinking generally. This ties closely with the subthemes of “Masking” and “All relationships carry expectations that alcohol helps manage”, as alcohol facilitates masking or unmasking in social settings which in turn would help foster connection with others. For some participants, this was particularly relevant to their work culture where they would try and fit in with colleagues.

#### Restrictive and repetitive behaviours and interests

“Restricted, repetitive patterns of behaviour, interests or activities” is another of the DSM-5-TR diagnostic criteria for autism.^27^ Some participants reported that drinking developed into a habit that formed a part of their routine and therefore became difficult to stop, in line with diagnostic criteria around repetitive patterns of behaviour. This is exemplified by one participant who was able to stop drinking after realising that the alcohol they routinely drank could be swapped out with the non-alcoholic equivalent - the alcohol itself was not that important, but the ritual that came with it was. Drinking also became automatically associated with certain activities for some participants, including meals, socialising with friends and watching television. Again, this highlights how autistic traits can interact with alcohol use and may be an important finding for autistic people struggling with their drinking who have not considered the influence of their routine. Moreover, an understanding of autistic features such as rigidity around routine was mentioned when discussing delivery preferences (in the “Recommendations for support” subtheme).

#### Sensory differences

Some participants stated that alcohol was useful for minimising the intensity of sound, smell and touch in social environments. This reduction in sensory input made it easier to be present and engage socially. For example, being in a loud, busy environment, or touched during sexual encounters become more manageable from a sensory perspective and therefore ties closely with the “social communication differences” subtheme. In contrast, taste sensitivity deterred some participants from drinking as they found it off-putting, suggesting that sensory differences do not always influence alcohol use the same way in autistic individuals, as would be expected in a highly heterogenous population.

#### Masking

“Symptoms were present in early childhood (but may not have become apparent until adulthood due to masking” is one of the criteria in the DSM-5-TR.^27^ Masking is the well-recognised phenomena where neurodivergent people, especially autistic people, employ strategies to appear neurotypical in order to avoid stigma or judgement for their authentic behavioural presentation.^28^ As previously mentioned, this subtheme relates more broadly to the “social communication differences” and “All relationships carry expectations that alcohol helps manage” subthemes. For some participants, alcohol made unmasking in social situations easier due to the perceived relaxation of those around them (as they therefore felt less judged by others while authentically expressing themselves). Others viewed alcohol as vehicle for masking, helping them to fit in and make friends by adhering what they perceived to be social norms (e.g. during work events). Several participants also highlighted a murky self-identity as a result of masking, with some of them still trying to figure out who they were ‘underneath’ the mask or without alcohol to help them unmask.

### Other autistic features

#### Risk aversion

Risk aversion is not specifically listed as a diagnostic criterion in the DSM-5-TR, but it has been described as a transdiagnostic construct that is a commonly reported consequence of uncertainty intolerance in autistic individuals.^27,29^ In contrast with many of the previous subthemes, risk aversion acted as a protective factor against hazardous drinking for some participants, preventing them from drinking altogether. People mentioned that the potential medical risks deterred them from heavy drinking, wanting to maintain awareness in social settings and not wanting to risk getting a hangover. Others however, still wanted to learn how to enjoy to alcohol occasionally in moderation, despite being apprehensive about the potential health risks. This is pertinent to the subtheme of “early alcohol education” as some participants felt that having a better understanding of alcohol from an earlier age would have helped them make more informed decisions around drinking.

#### Coping using alcohol

The second theme we generated was “Coping using alcohol”. Many participants described using alcohol to cope with social expectations from partners, colleagues, children and friends. This was reflected in the subtheme “All relationships carry expectations that alcohol helps manage”. Participants used alcohol as a form of escape from these responsibilities to be able to reset and show up as expected, so we captured this using the subtheme “Drinking is a form of escape and a chance to reset”. Additionally, for some, drinking was also a form of self-medication for various co-occurring difficulties. Our final subtheme was therefore titled “Drinking to self-medicate”.

### All relationships carry expectations that alcohol helps manage

Some participants reported that drinking alcohol allowed them to stave off potential burnout from having to perform at their job by giving them an outlet or relief. Stressors at work included having to mask, engage with colleagues, sensory input at the office, ableism and increased general responsibility over time that alcohol allowed them to cope with. Work culture was also a reason for drinking, as that was what colleagues would do to socialise and they wanted to fit in with the team. As previously mentioned, experiences such as these at work relate directly to the subthemes of “social communication differences” and “masking”. Further, living with partners and/or children was also a source of stress and exhaustion for some participants. One explained that they were unsure whether they would have been a parent had they known they were autistic because of how much time alone they required. Further, many participants found it easier to socialise when drinking, even with friends.

### Drinking is a form of escape and a chance to reset

Several participants used alcohol to disassociate, wind down and numb the mind at the end of the day as a form of escapism from negative emotions. For some, doing this was what allowed them to reset before tackling the next day and reflected attempts to manage expectations in relationships (as explained in the previous subtheme), particularly in parenthood and full-time employment.

### Drinking to self-medicate

In similar vein, many participants reported self-medicating with alcohol to manage a variety of difficulties. These included stress, anxiety, muscular tension, overstimulation, suicidal ideation, exhaustion, unhappiness, emotional pain, desperation, insomnia and trauma. One participant even described their drinking as a form of self-harm. Evidently many participants shared the same motivation (to manage a difficult feeling by drinking) for many different issues. Some participants also commented on how they self-medicated prior to receiving their autism diagnosis for issues such as anxiety that they were unaware were linked to their autism.

**Table 2.** Quotes illustrating subthemes of “Managing expectations and coping using alcohol”.

| Subtheme | Quote | Participant # |
| --- | --- | --- |
| ***All relationships carry expectations that alcohol helps manage*** | “ I was living with someone, had kids, had a job. I didn’t realise how much stress those things were putting on me” | 12 |
|  | “ So, yes, I think every day, apart from Saturday, was a drinking day. I think it was all about the work, you know... having to sit next to somebody, having to answer the telephone, engage with people, just having the general noise of people clattering on computers or phones ringing, or people chatting or laughing.” | 13 |
| ***Drinking is a form of escape and a chance to reset*** | “ I didn’t realise how much I needed [to drink] to be able to get up the following day and perform like a neurotypical.” | 11 |
|  | “It was always a form of emotional experience, relaxation, and escape” | 6 |
| ***Drinking to self-medicate*** | “ You could see that as a positive, but obviously, there are negative connotations. There was also an element of numbing. I’ve always dealt with very high levels of anxiety, which, until very recently, I didn’t realise were linked to autism. I’ve dealt with phenomenal levels of anxiety since my early to mid-teens. The amount of alcohol I was drinking helped numb or suppress that, which felt good at the time. But, obviously, with hindsight, I can see how unhealthy that behaviour was.” | 18 |
|  | “So, by having a couple of glasses of wine in the evening, it would help me wind down quicker and get into bed, where I’d fall asleep more easily.” | 14 |
|  | “...it slows down my thinking, self-consciousness, and self-criticism.” | 1 |

#### Recommendations for support

The final theme we generated was “Recommendations for support”. The inaccessibility of existing services was of particular concern (reflected in the subtheme “Current services are inaccessible”). Further, some participants expressed a desire for alternatives to group settings, which led us to create the subtheme “Delivery preferences”. In addition to this, participants expressed the desire for services to be autistic led (or at the very least have an informed understanding of autism generally). This was encompassed by the subtheme “Autism at the heart of services”. Finally, participants stated they would like to see more education about alcohol in schools, which we denoted with the subtheme “Early alcohol education”.

### Current services are inaccessible

The main service discussed by participants was group meetings, particularly Alcoholics Anonymous (AA). Some participants had accessed AA themselves, while others had not but commented on the group structure. Communicating in a group setting was the main reason for inaccessibility, but unstructured breaks and absorbing the emotional difficulties of other group members were also off-putting. These elements combined to mean that those who had accessed services found them inaccessible and had discouraged several participants from accessing help for their drinking, despite a desire for support. For some participants this formed the foundation for the next subtheme (“delivery preferences”).

### Delivery preferences

With existing services often felt to be inaccessible, some participants felt that the option of one-on-one support would be more appropriate, with the flexibility to attend either online or in person (depending on personal preference). They felt that this would be more effective because it would mean circumnavigating the issues around inaccessibility explained in the previous subtheme, and ties closely with the next subtheme (“autism as the heart of services”).

### Autism at the heart of services

Many participants expressed a desire for services to understand autistic ways of thinking, and to validate their experiences generally (which follows on from “delivery preferences”). For example, some participants highlighted key autistic features they felt should be considered in a service context, such as the idea that sobriety can inhibit masking and subsequently induce social anxiety. One participant also touched upon the need for consistency, suggesting that it would have been helpful to know that alcohol could be swapped for a non-alcoholic equivalent as part of their routine. Another also highlighted that it is important for services to understand that autistic people can take time to change generally. Further, participants highlighted that service providers should be understanding in group settings if autistic people need to be alone and therefore do not attend consistently. However, they also highlighted the importance of balancing that with the risk of isolation precipitating a relapse. Some people did not mind whether service leaders were autistic or not, providing staff were autism informed. Other participants preferred the idea of autistic service leaders as they felt they would be more likely to actually listen to autistic service users, understand their experiences, and have innovative ideas for support.

### Early alcohol education

Some participants explained that an aversion for risk deterred them from drinking once they were aware of the potential health and social repercussions (hence the link to the subtheme “risk aversion”). Some expressed a desire to see more education in schools around alcohol to make it less of a novelty and the potential risks and general consumption guidelines clearer. Those who raised this point felt that having this knowledge from before they first started drinking would have discouraged them from ever reaching hazardous levels, providing a lifelong protective function.

**Table 3.** Quotes illustrating subthemes of “Recommendations for support”.

| Subtheme | Quote | Participant # |
| --- | --- | --- |
| **Current services are inaccessible** | "I can't imagine what it would be like for an autistic person going through that kind of situation with so many people in AA meetings. I just don't think they're set up for someone with autism." | 14 |
|  | "It's not that I didn't want to engage, but I didn't like having that break in the middle." | 11 |
| **Delivery preferences** | "Oh, yeah, certainly the option of one-to-one. Maybe, for those who feel comfortable, small groups." | 17 |
|  | "... a remote access option, might be useful. Although, obviously, I appreciate that some people don't like talking on computers and so on." |  |
| **Autism at the heart of services** | "The major adjustment most services should have is involving more autistic individuals with alcohol lived experience because they would have a higher likelihood of being listened to and [Someone autistic] would understand the plight better" | 10 |
|  | "I can't comment on existing services because I don't know what they are, but they should include an understanding that, first of all, autistic people are different from each other, not just from neurotypicals... As a generalisation, autistic people can take longer to change their trajectory. So even if their reason for drinking disappears, they may not be able to stop... If someone dealing with an alcohol issue doesn't understand autism, how can they help? They could unintentionally make things worse because they wouldn't understand the nuances that underlie everything." | 8 |
| **Early alcohol education** | "Just understanding the drinking, like, "Oh, this drink has so many units in it, and this is how many you're allowed a day." I didn't know any of that. I was just drinking and drinking and drinking. It was only when I got to maybe 16, 17, or 18 that I started to become aware of, "Oh, you're only allowed this many units if you drive, and that's what's safe." Then you're like, "Oh my gosh, I was having 20-30 units in one night." That's loads more than you should have if you're in charge of a car, you know? I was learning all these things a bit too late. I mean, I may not have listened to them, but I can't make decisions if I don't have the information." | 3 |

## 6.4 Discussion

In this study, eighteen autistic adults in the UK described their experiences around alcohol use. Novel insights were uncovered and these varied between participants, shedding further light on some of the nuances around alcohol use in the autistic community while extending the limited research on this topic. Three main themes were generated, however. These encompassed the ways in autistic features influenced alcohol use, how alcohol could be a coping tool, and a discussion about existing services and potential adaptations to support autistic people. At present this area remains under-researched, and

From these findings it is apparent that being autistic can directly influence an individual’s relationship with alcohol (though there is obviously variation within this). As reflected in the literature, participants reported using alcohol to facilitate social interactions by increasing confidence, comfort and inclusion.^13,14^ Participants elaborated on these findings by attributing these easier social interactions to specific changes in both themselves and others, including increased mutual understanding and honesty. However, social communication differences are not exclusive to autism but can also appear with other psychiatric differences including ADHD and schizophrenia.^30^ Alcohol services therefore may find it beneficial to consider their role in the initiation and maintenance of alcohol use in autistic and non-autistic individuals alike.

Nearly all participants in our study were late diagnosed and were predominantly women. This is reflective of late autistic diagnoses generally, with autistic women being less likely to be diagnosed in childhood than men.^27,31^ Some particularly interesting commentary was captured from female participants, which may have important implications for supporting autistic women. Some of the female participants discussed the exhaustion they felt from masking when being a colleague, partner and parent simultaneously (without realising that is what they were doing at the time as this was pre-diagnosis). This algins with existing research around masking in autistic women, where concealing autistic traits has been linked to stress, anxiety, depression and exhaustion and may delay diagnosis.^32–34^

These social responsibilities denied participants the time alone required to recharge and as a result they turned to alcohol to try and meet this need so they could function as expected. Qualitatively, autistic individuals have reported a need (rather than want) to have time alone so that they then have the energy required to connect with others.^35^ Based on our findings, is therefore feasible that social expectations and a lack of self-understanding pre-diagnosis may deny autistic women from the lone time they need to recharge. In turn, these may leave autistic women more at risk of developing an alcohol dependence than previously thought as they try to cope with the pressure of social expectations and consequent lack of space to rest.

Moreover, self-medication was another motivation for participants when drinking that aligned with existing findings – specifically for overstimulation and overwhelm - which are consistently identified as key wellbeing issues for autistic adults. This same motivation has been reported by other autistic individuals in a recent qualitative study.^36^ Additional difficulties that participants managed with alcohol included stress, anxiety, muscular tension, suicidal ideation, exhaustion, unhappiness, emotional pain, desperation, insomnia and trauma (many of which are common in autistic individuals generally).^37–39^ Given the extent of issues that participants have reported using alcohol to cope, and the higher co-occurrences of such issues in autistic people compared to non-autistic people, it is unsurprising that there is emerging evidence to suggest an increased risk of hazardous drinking in members of the community.^10,40,41^

Finally, these findings show us why current services may not work for some autistic people and what can be done about it. Nearly all participants felt that autistic people needed tailored support with hazardous drinking, with concerns about the inaccessibility of existing services (particularly in relation to group settings). Participants expressed a preference for alternatives such as one-to-one in person and/or online support and autism informed services. These suggestions are in line with recommendations from the community for support with a range of issues, such as Relationship and Sex Education (RSE), eating disorders (EDs), and mental health conditions.^42–44^ An example where autism-led services have been implemented successfully is the Pathway for Eating disorders and Autism developed from Clinical Experience (PEACE) model.^45^ The PEACE model is an evidence-based framework centred around the adaptation of existing eating disorder services to support members of the autistic community, with promising results surrounding implementation across Buckinghamshire, Oxfordshire and Berkshire (BOB).^46^ Core principes of PEACE include recognition of the observed overlap between autism and EDs, educating staff, autism assessment upon intake into ED services, adapting treatment delivery and community engagement.^45^

There are existing guidelines (‘Clinical guidelines for alcohol treatment’) published by The Department of Health and Social Care where detailed recommendations are given under section 25.11 around adjustments for neurodivergent individuals accessing alcohol services. These are similar to the PEACE model; in summary, they include having adequate staff training, supervision and support and an awareness of co-occurring mental health issues (e.g. suicide risk), individuals being potentially undiagnosed. Other recommendations include being autism-led, reducing sensory input in service areas where possible, communicating information accessibly (and adapting where necessary) and involving loved ones and/or other services for support. Finally, service users should be encouraged to engage in structured recreational activity, receive tailored employment support if required and expertise sought when providing pharmacological interventions. However, there has yet to be a formal implementation of these recommendations that can then be assessed as there has been with the PEACE model (and no funding allocated to do so). It is possible that following a similar process to trial the adaptation of an existing alcohol service based on these guidelines could yield similarly promising results as the PREACE implementation project across BOB.^43^

Following the results of this paper, a possible addendum to the guidelines mentioned could be for clinicians to be mindful of the role alcohol can play in the lives of autistic women and the influence of delayed diagnosis. Existing services such as Alcoholics Anonymous (AA) may also find it useful to make adaptations to improve the accessibility of group meetings for autistic individuals, such as ensuring the availability of online meetings and/or one-to-one sessions where possible. Further, participants wished to see greater early education in schools, to help children make more informed decisions around drinking as they get older. As such, an additional recommendation for guidelines could be the implementation of early education around alcohol geared towards to pupils receiving SEND (Special Educational Needs and Disabilities) support (and pupils generally to reach undiagnosed individuals). We appreciate that such recommendations may be difficult to implement when resources are constrained but hope they remain valuable nonetheless for service providers. Without appropriate support, members of the autistic community may continue to struggle with their drinking alone when this could be preventable.

### Strengths and limitations

This study has several strengths. To our knowledge, it is the first qualitative study to explore alcohol use in autistic adults in a non-clinical sample. Our findings have added nuance around the motivations for drinking alcohol among members of the autistic community. These include changes in demeanour that facilitate social interactions, self-medication of a plethora of co-occurring difficulties, and managing the pressure of social expectations (particularly for working mothers and those in long-term relationships). Finally, participants have given specific preferences for therapeutic interventions, supporting the clinical utility of findings.

There also some notable limitations of this study. Firstly, the sample is predominantly white and female. One on hand it is positive to have research that focuses on autistic women given the historic exclusion of this group from autism research (partially due to the historic assumption that women ‘could not be’ autistic)^47^ However, the experiences of other genders and non-white individuals are likely to have intricacies not reported in this paper.^48,49^ Secondly, the UK is notorious for its binge drinking culture and has some of the highest alcohol use per person globally.^50^ It is therefore possible that individuals based in countries that have other attitudes towards alcohol would report different experiences (for example in predominantly Muslim countries where alcohol use is discouraged as part of the faith)^51^. Evidently, there is more to uncover in such a diverse community, and there must be some hesitation about generalisation of findings to autistic people worldwide.

### Conclusions

In conclusion, our participants highlighted the ways in which autistic features can influence alcohol use and shed light on some of the mechanics that underlie the facilitation of social interactions for autistic people when drinking. Further, we have seen the range of struggles that autistic people face which can drive them to self-medicate with alcohol. Participants have also demonstrated a desire for autistic-led or autism-informed services, as well as alternative options to group settings and a desire for early education around alcohol. The level of interest expressed by members of the community in taking part in this study demonstrates a clear desire for further research in this area. Service providers and policy makers would likely benefit from taking the findings from this paper into consideration and any similar research in the future involving other demographics within the autistic community.

## Data Availability

All data produced in the present study are available upon reasonable request to the authors

## Acknowledgements

The authors would like to express their gratitude to the participants and advisory group for engaging in this research, providing invaluable insights that we hope will be of help to the autistic community. We would also like to thank Autistica for their support.

